# Normative Auditory Nerve Physiology Reveals Myelin-Associated Changes in Neural Aging

**DOI:** 10.64898/2026.09.11.26362834

**Authors:** Kelly C. Harris, Shelby A. Payne, James W. Dias, Abigail Lawson, Courtney Hall, Carolyn M. McClaskey, Judy R. Dubno, Hainan Lang

**Affiliations:** Medical University of South Carolina: Department of Otolaryngology-Head and Neck Surgery; Department of Pathology and Laboratory Medicine. Medical University of South Carolina, Walton Research Building, 39 Sabin Street, Charleston, SC 29425; Ralph H. Johnson Department of Veterans Affairs Medical Center, 109 Bee Street, Charleston, SC 29401

## Abstract

Current physiological measures of auditory nerve function reveal neural dysfunction but provide limited insight into its underlying mechanisms. Myelin degeneration is an important feature of neural aging, yet identifying myelin-associated auditory nerve dysfunction in living humans remains challenging. We developed a normative physiological approach by modeling the relationship between auditory nerve response amplitude and neural synchrony in younger adults and quantifying deviations from this relationship in older adults, including both females and males. The strong coupling between amplitude and synchrony observed in younger adults was markedly weakened with aging. In older adults, poorer-than-expected synchrony relative to response amplitude was associated with lower fractional anisotropy and higher radial and mean diffusivity in the auditory nerve, a pattern consistent with reduced myelin integrity. Diffusion MRI and histology from two human temporal bone specimens showed corresponding differences in diffusion measures and myelin structure, providing preliminary histological support for this interpretation. A descriptive partition of older adults by residual synchrony further revealed that the relationship between audiometric thresholds and self-reported hearing difficulty differed according to physiological grouping. Hearing difficulty increased more steeply with worsening thresholds among those with poorer-than-expected synchrony. Together, these findings suggest that deviations from normative auditory nerve physiology capture microstructurally and behaviorally relevant variation in neural aging. This approach provides a foundation for developing physiological biomarkers that may ultimately help distinguish mechanisms of auditory nerve dysfunction and guide mechanism-targeted interventions.

**Significance Statement:** Conventional hearing tests do not capture age-related changes in auditory nerve function or distinguish the physiological processes contributing to neural dysfunction. We introduce an approach that evaluates neural synchrony relative to auditory nerve response amplitude, moving beyond identifying reduced neural function toward detecting disproportionate impairments in temporal precision. Deviations from the relationship established in younger adults were associated with diffusion MRI measures of auditory nerve microstructure, supported by illustrative MRI–histological findings from human temporal bones. This physiological measure also helped explain self-reported hearing difficulty. By linking electrophysiology, neuroimaging, human temporal bone pathology, and patient-reported outcomes, these findings establish a framework for developing clinically accessible measures that may help distinguish mechanisms of auditory nerve dysfunction, including myelin-associated changes.

## Introduction

Age-related hearing loss is among the most common chronic conditions affecting older adults and has consequences beyond reduced audibility (Gates and Mills, 2005). Yet elevated audiometric thresholds explain only a portion of variability in communication difficulties (Dubno et al., 1984). Older adults with similar audiometric thresholds may report markedly different communication abilities, suggesting that conventional audiometric measures fail to capture important age-related changes elsewhere in the auditory system.

The auditory nerve (AN) is particularly vulnerable to aging, with degeneration occurring before substantial sensory hair-cell loss (Makary et al., 2011; Xing et al., 2012; Wu et al., 2019). Animal studies have identified synaptic loss, myelin degeneration, and axonal degeneration as contributors to AN dysfunction (Schmiedt et al., 2002; Xing et al., 2012; Kujawa and Liberman, 2015; Wan and Corfas, 2017; Panganiban et al., 2018; Hickman et al., 2021; Suthakar and Liberman, 2021; Panganiban et al., 2022). Consistent with these findings, human electrophysiological studies demonstrate age-related reductions in AN response amplitude and neural synchrony, even among older adults with clinically normal hearing thresholds (Harris et al., 2018; Harris et al., 2021; Rumschlag et al., 2022; Dias et al., 2024). Moreover, reduced neural synchrony is associated with poorer speech recognition in noise and altered cortical processing, suggesting that peripheral AN dysfunction may initiate broader changes throughout the central auditory system (Seki and Eggermont, 2003; Kotak et al., 2005; Sanes and Bao, 2009; Dong et al., 2010; Popescu and Polley, 2010; Engineer et al., 2011; Kraus et al., 2011; Salvi et al., 2016; Parthasarathy et al., 2019; Harris et al., 2022). Although these physiological measures provide evidence of AN dysfunction, they offer limited insight into whether the underlying pathology involves synaptic loss, myelin degeneration, or other forms of neural injury. Distinguishing among these cellular and sub-cellular mechanisms is important because they may require different target therapeutic approaches.

Our previous work demonstrated that AN physiology is not fully characterized by response amplitude alone (Harris et al., 2018; Harris et al., 2021). In younger adults with normal hearing, larger response amplitudes are accompanied by greater neural synchrony, reflecting the normal coupling between the number of responding fibers and the precision of their temporal firing (Harris et al., 2021). Re-examination of these data revealed that this relationship was markedly weaker in older adults. This finding suggested that aging disrupts the normal coupling between AN response amplitude and neural synchrony, raising the possibility that some older adults exhibit disproportionately poor neural synchrony relative to their AN response. This approach parallels a broader shift in neurodegenerative disease research from examining biomarkers individually or as group-level differences toward identifying individual deviations from normative biological relationships (Verdi et al., 2023; Verdi et al., 2024). Applying a similar framework to AN aging may reveal biologically meaningful forms of neural dysfunction that are not apparent from either physiological measure alone.

Our previous studies in mouse models demonstrated that neural synchrony is sensitive to AN myelin integrity (Panganiban et al., 2022). Myelin is also an important therapeutic target because it retains the capacity for self-repair and remodeling throughout adulthood (Franklin and Ffrench-Constant, 2017; Bonetto et al., 2020). However, physiological measures capable of distinguishing myelin-associated dysfunction from other forms of AN pathology have not been established in living humans. Advances in remyelination therapies underscore the need for biomarkers of neural dysfunction. We hypothesized that deviations in neural synchrony from that expected based on AN response amplitude identify a physiological pattern of AN aging associated with AN myelin dysfunction/degeneration. We established a normative model relating AN response amplitude to neural synchrony in younger adults and quantified deviations from this relationship in older adults. We then tested whether older adults assigned to the resulting physiological phenotypes differed in AN diffusion MRI measures associated with myelin integrity and examined correspondence between these diffusion measures and myelin pathology in human temporal bone specimens. Finally, we tested whether physiological phenotype explained differences in self-reported hearing difficulties beyond conventional audiometric thresholds.

## Materials and Methods

### Participants

A total of 182 adults participated in the study, including 69 younger adults (18–30 years; 49 female) and 113 older adults (54–83 years; 77 female). Younger adults had normal hearing, defined as air-conduction thresholds ≤20 dB HL from 0.25 kHz to 8 kHz. Older adults exhibited a broad range of hearing thresholds, ranging from normal hearing to moderate sensorineural hearing loss. Air-and bone-conduction thresholds were obtained using a Madsen audiometer calibrated according to ANSI standards. Hearing thresholds were summarized using both the four-frequency pure-tone average (PTA4; 0.5, 1, 2, and 4 kHz) and the high-frequency pure-tone average (PTAHF; 2, 4, and 8 kHz).

Inclusion criteria included English as a primary language and a Mini-Mental State Examination score ≥27. Exclusion criteria included conductive hearing loss, active otologic disease, history of significant head trauma or seizures, neurological disorders affecting the central nervous system, and contraindications to MRI.

All participants underwent AN electrophysiological testing. Results from younger adults were used to establish the normative relationship between AN response amplitudes and neural synchrony. Based on those results, two physiological AN phenotypes were identified within the older adult cohort, as described below. Diffusion MRI and self-reported hearing difficulty measures were collected as part of ongoing studies and were therefore available for subsets of participants. Human temporal bone MRI and histology were obtained from separate postmortem specimens, as described below. Diffusion MRI analyses included 18 younger adults and 47 older adults, whereas self-reported hearing difficulty measures were available for 31 younger adults and 74 older adults. Eleven younger adults and 34 older adults had both diffusion MRI and RHHI data. Of the 182 participants, 176 (67 younger and 109 older) had usable CAP amplitude and PLV measurements. Statistical analyses were performed using all available data for each outcome measure (Table 2). All participants provided written informed consent under protocols approved by the Medical University of South Carolina (MUSC) Institutional Review Board.

### AN Electrophysiology

Compound action potentials (CAPs) were recorded using a tympanic membrane (TM) electrode as previously reported (Harris et al., 2018; Harris et al., 2021). CAPs were elicited using 100-µs rectangular clicks of alternating polarity presented monaurally to the right ear through ER3C insert earphones (Etymotic Research) at 110 dB peak sound pressure level. Stimulus generation and presentation were controlled using RPvdsEx software (Tucker-Davis Technologies). Responses were recorded in two blocks of 1100 stimulus presentations (550 of each polarity).

CAPs were recorded using a tympanic membrane (TM) electrode (Sanibel Supply) placed on the right TM, a contralateral mastoid reference electrode, and a low-forehead ground electrode. Auditory brainstem responses were simultaneously recorded using a high-forehead active electrode and ipsilateral mastoid reference to assist with identification of the CAP N1 response. Signals were amplified using a Neuroscan SynAmpsRT amplifier operating in AC mode with a gain of 2010× and digitized at 20 kHz using Tucker-Davis Technologies hardware. Recordings were obtained in an electrically and acoustically shielded booth while participants rested quietly and were permitted to sleep.

Continuous recordings were analyzed offline in MATLAB (MathWorks, Natick, MA) using EEGLAB and ERPLAB. Signals were band-pass filtered from 150 to 3000 Hz. Stimulus triggers were corrected for the 1.0-ms earphone delay and 0.6-ms delay introduced by the Tucker-Davis digital-to-analog converter. Data were epoched from −2 to 10 ms and baseline corrected using the −2 to 0 ms prestimulus interval. Trials containing voltage excursions greater than ±45 µV were rejected automatically and subsequently reviewed by visual inspection. Remaining trials were averaged to generate the CAP waveform. N1 peak amplitude was measured relative to the prestimulus baseline using custom MATLAB routines within ERPLAB. Peak identification was independently verified by two experienced reviewers and evaluated across repeated analyses for consistency.

Neural synchrony was quantified using phase-locking value (PLV), an estimate of intertrial phase consistency derived from individual electrophysiological responses. PLV ranges from 0, indicating no phase consistency across trials, to 1, indicating perfect phase consistency. Time-frequency decomposition was performed using the EEGLAB newtimef() function with Hanning-window fast Fourier transform tapers. Analyses included 16 linearly spaced frequencies from 625 to 2500 Hz using a 64-sample analysis window and a pad ratio of 2. A single PLV value was obtained for each participant by identifying the peak PLV occurring within a 2-ms window surrounding the CAP N1 response.

To assess the influence of recording quality on neural synchrony estimates, additional measures were calculated from cleaned single-trial CAP recordings. Prestimulus phase consistency was estimated from the 2-ms interval preceding acoustic stimulus arrival using Fourier analysis restricted to prestimulus samples (500–2500 Hz, 500-Hz increments). We calculated prestimulus PLV and pairwise phase consistency (PPC), a bias-corrected measure of squared phase coherence, along with prestimulus root-mean-square (RMS) voltage and accepted trial counts. Run-level measures were averaged within participants. The prestimulus estimator used a shorter window than the evoked PLV estimator and was intended as a complementary recording-quality control rather than an identical measure of the evoked PLV noise floor.

### AN Diffusion MRI

Diffusion MRI was acquired on a Siemens Prisma 3T MRI scanner using a 64-channel head/neck coil and a 128-direction monopolar diffusion-weighted sequence (1.5-mm isotropic voxels, *b* = 700 s/mm², TR = 6000 ms, TE = 50 ms), together with a reverse phase-encoded acquisition for susceptibility distortion correction. Diffusion preprocessing was performed using FSL together with Diffusional Kurtosis Estimator software and custom MATLAB scripts. Images underwent susceptibility distortion correction, and motion correction, tensor estimation, and calculation of fractional anisotropy (FA), mean diffusivity (MD), and radial diffusivity (RD).

Because of the small size of the AN relative to the spatial resolution of diffusion MRI, regions of interest (ROIs) were conservatively delineated to minimize partial-volume contamination from surrounding cerebrospinal fluid and adjacent neural structures. Two experienced raters used FA and MD maps, diffusion-weighted images, and anatomical landmarks, including the cochlea, to identify portions of the right auditory nerve with high confidence. Rather than imposing a fixed anatomical extent, segmentation prioritized anatomical specificity, and consensus masks were generated before extracting mean diffusion measures. Inter-rater reliability was evaluated in 25 participants using independently delineated ROIs and intraclass correlation coefficients for ROI size and diffusion measures. Sensitivity analyses examined whether the physiological–diffusion associations were influenced by ROI size.

### Human Temporal Bone MRI and Histology

Two human temporal bone specimens were obtained from the MUSC Research Program temporal bone archive. Specimens were obtained from a male donor in his 50s (specimen 1) and female donor in her 80s (specimen 2), with post-mortem intervals <10 h. Written informed consent for temporal bone collection was obtained from next of kin in accordance with South Carolina laws and regulations. Temporal bone research was approved by the MUSC Institutional Review Board as not human subjects research.

The procedures for the collection and process of the human temporal bones were performed as previously described (Xing et al., 2012). After removal of the temporal bone from the skull, scalar perfusion was performed with 4% paraformaldehyde, followed by immersion fixation for at least 48 h at 4°C. Prior to histological processing, temporal bones were embedded in 6% agar and imaged on a Bruker 7T small-bore MRI system. High-resolution T2-weighted, diffusion-weighted, and quantitative T1 images were acquired. Diffusion imaging protocols were optimized for postmortem tissue to maximize signal-to-noise ratio while preserving the spatial resolution required to visualize the AN. AN ROIs included the nerve visible within the temporal bone specimen, extending from the cochlea through the available nerve to the proximal end of the specimen. Diffusion tensor metrics were calculated from manually defined AN ROIs using Diffusional Kurtosis Estimator software and custom MATLAB scripts adapted for postmortem tissue.

Following MRI acquisition, the portion of the AN previously examined using MRI was dissected from the internal auditory meatus and sectioned transversely at 12 µm. Sections were stained with FluoroMyelin™ Fluorescent Myelin Stain to label myelin-associated structures and propidium iodide to label cell nuclei as previously described (Xing et al., 2012; Payne et al., 2026). The images were collected and analyzed using a Zeiss LSM 880 NLO with Airyscan and ZEN acquisition software.

Myelin thickness, axon diameter, and g-ratio were measured manually using Fiji ImageJ. Fibers were included only when the perimeter of both the axon and myelin sheath could be clearly distinguished. G-ratio was calculated as the ratio of axon diameter to myelinated fiber diameter. Histological observations were compared directly with diffusion MRI measures obtained from the same temporal bone specimens.

### Physiological AN Phenotyping

Prior to analysis, CAP amplitude and PLV were standardized (z-scored) to place the two physiological measures on a common scale. A linear regression relating standardized CAP amplitude to standardized PLV was fit using data from younger adults to establish the normative physiological relationship between AN response amplitude and neural synchrony. This model was then applied to each older adult to determine the PLV predicted based on their CAP amplitude. Residual neural synchrony was calculated as observed PLV minus predicted PLV, with increasingly negative residuals indicating poorer neural synchrony than expected based on the AN response amplitude.

To identify physiological subgroups without incorporating MRI, audiometric, or self-report outcomes into the classification, residual neural synchrony values were classified using unsupervised k-means (Hartigan and Wong, 1979). The optimal number of clusters was evaluated using average silhouette width and gap statistic analyses. Both approaches supported a two-group descriptive partition, yielding poorer-than-expected and as-expected-or-better synchrony phenotypes relative to younger normative physiology. Because clustering a single continuous residual measure does not establish distinct underlying populations, residual synchrony was also analyzed continuously, and the cluster labels were interpreted as descriptive groupings rather than discrete biological subtypes. Stability of the two-cluster classification was evaluated using 1,000 bootstrap resamples of older adults. For each resample, k-means clustering was repeated, and the resulting cluster centers were used to classify the original participants. Agreement with the original classification was quantified using the adjusted Rand index.

### Self-reported hearing difficulty

Communication difficulty was assessed using the Revised Hearing Handicap Inventory (RHHI), a validated patient-reported outcome measure of hearing-related psychosocial health. Higher scores indicate greater perceived hearing difficulty (Cassarly et al., 2020).

## Statistical Analysis

Statistical analyses were performed in R version 4.3.0 (R Foundation for Statistical Computing, Vienna, Austria) and MATLAB R2024b (MathWorks, Natick, MA, USA). All statistical tests were two-sided, with statistical significance defined as *p* < 0.05. Model assumptions, including normality and homoscedasticity of residuals, were evaluated by inspection of diagnostic plots.

Age-related differences in CAP amplitude and PLV were evaluated using linear regression. To determine whether the relationship between AN response amplitude and neural synchrony differed with age, PLV was modeled as a function of CAP amplitude, age group, and their interaction. Within younger adults, the association between CAP amplitude and PLV was quantified using Pearson correlation and linear regression to establish the normative physiological relationship, as described above. To assess whether age-group differences in CAP amplitude–PLV relationships were influenced by differences in the range of CAP amplitudes, the regression analysis was repeated using observations within the overlapping amplitude range of the two age groups. Sensitivity to extreme observations was evaluated by excluding the two younger adults with the largest-magnitude CAP responses.

The distribution of residual neural synchrony was examined using Gaussian mixture modeling, with the Bayesian information criterion used to compare models with different numbers of components. The stability of the two-cluster classification was evaluated using bootstrap resampling, as described above. Additional sensitivity analyses examined changes in phenotype assignment when residuals were derived using robust regression or after excluding the two younger adults with the largest-magnitude CAP responses from the normative model.

Within older adults, differences in age, PTA4, and PTAHF between physiological phenotypes were evaluated using linear regression. Associations between physiological phenotype and AN diffusion MRI measures (FA, MD, and RD) were evaluated using linear regression adjusted for age and PTA4. Unadjusted between-phenotype effect sizes were quantified using Hedges’ *g*. Continuous associations between residual neural synchrony and diffusion MRI measures were also examined using linear regression adjusted for age and PTA4. Because FA, MD, and RD were highly correlated, principal component (PC) analysis was used to derive a composite measure of AN microstructure (PC1).

To compare the information provided by individual physiological measures and the normative residual, complete-case linear models examined AN diffusion PC1 in 47 older adults, with age and PTA4 entered first and CAP amplitude, raw PLV, or residual neural synchrony added separately. Model improvement was evaluated using changes in explained variance and nested-model tests. An additional nested-model comparison evaluated whether CAP amplitude improved model fit beyond raw PLV. Sensitivity analyses examined the association between residual neural synchrony and PC1 when the normative relationship was estimated using robust regression or after excluding the two extreme younger-adult observations. Additional models included AN ROI size as a covariate.

Prestimulus PLV, PPC, RMS voltage, and accepted trial counts were compared between age groups and between older-adult physiological phenotypes using Welch’s *t* tests. Spearman correlations were used to examine associations of prestimulus PPC, RMS voltage, and accepted trial counts with evoked PLV and residual neural synchrony within older adults. These analyses evaluated whether the physiological measures covaried with prestimulus recording-quality indices.

Associations between physiological phenotype, audiometric thresholds, and self-reported hearing difficulty were evaluated using hierarchical linear regression. PTA4 was entered first, followed by physiological phenotype and the PTA4 × phenotype interaction. Improvement in model fit at each step was evaluated using the change in explained variance (ΔR²) and nested-model *F* tests. Additional models compared interactions between PTA4 and CAP amplitude, raw PLV, or residual neural synchrony in the same 70 older adults.

Analyses were performed using all available data for each outcome; therefore, sample sizes differed across electrophysiological, diffusion MRI, and RHHI analyses as described above. To assess potential sex-related confounding, we compared sex distribution across physiological phenotypes using Fisher’s exact test and conducted sensitivity analyses incorporating sex as a covariate in the primary electrophysiological and diffusion MRI models.

## Results

### 1. Results from younger adults establish the normative relationship between AN amplitude and neural synchrony

Compared with results from younger adults, and consistent with our prior studies, older adults exhibited significantly smaller CAP amplitudes (*p*= 0.012) and lower PLVs (*p* = 0.019) than younger adults (Fig. 1A). Moreover, the association between CAP amplitude and PLV differed significantly between age groups (age × amplitude interaction, *p <0.001*) (Fig. 1). CAP amplitude was strongly associated with PLV in younger adults (*r* = −0.85, *p* < 0.001), establishing the normative relationship between AN response amplitude and neural synchrony. This normative model was subsequently used to quantify deviations between CAP amplitude and neural synchrony among older adults.

**Figure 1.**
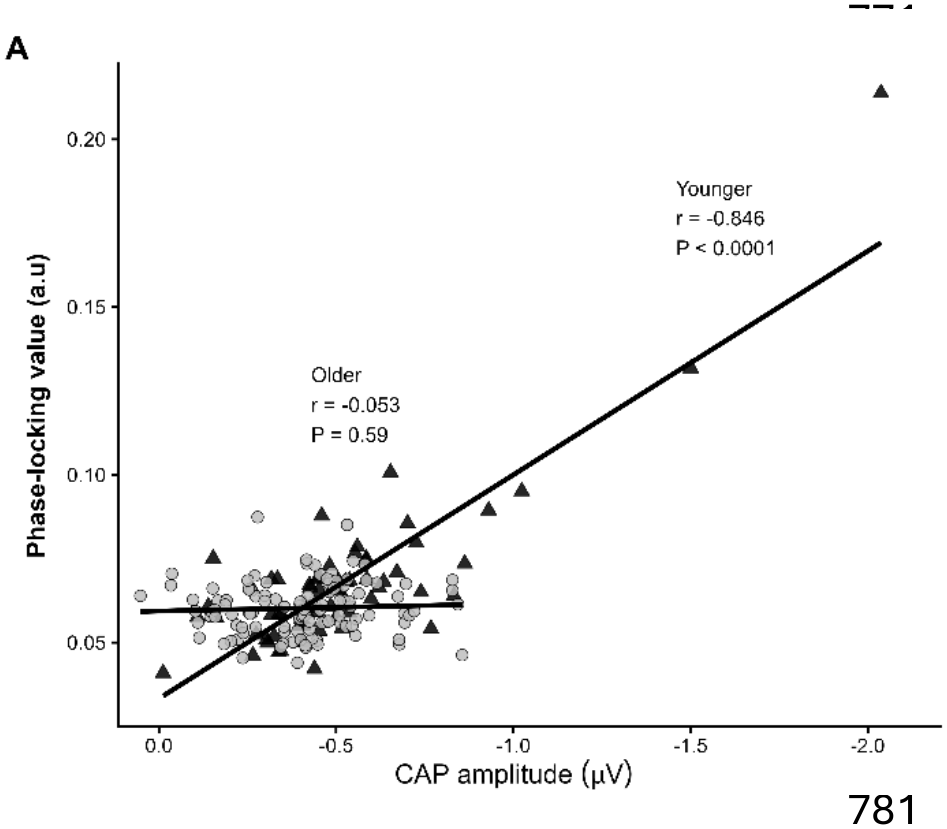
Age modifies the association between auditory nerve (AN) response amplitude and neural synchrony. CAP amplitude is strongly associated with phase-locking value (PLV) in younger adults (black triangles), establishing the normative relationship between AN response amplitude and neural synchrony. In contrast, this relationship is absent in older adults (gray circles), despite generally similar ranges of CAP amplitudes. Solid lines represent least-squares linear regressions for each age group.

To assess potential effects of recording quality, we compared prestimulus phase coherence, RMS voltage, and accepted trial counts between age groups. Prestimulus PLV did not differ significantly between younger and older adults (0.026 vs. 0.027, *p* = 0.390), nor did PPC (0.000007 vs. 0.000019, *p* = 0.787), RMS voltage (8.68 vs. 9.02 µV, *p* = 0.089), or accepted trial count (1219 vs. 1201, *p* = 0.576). Across the matched age-group sample, prestimulus PPC was not significantly associated with evoked PLV (Spearman ρ = 0.062, *p* = 0.426). Second, to determine whether the weaker relationship between CAP amplitude and PLV in older adults reflected their more restricted range of CAP amplitudes (SD = 0.188 µV vs. 0.294 µV in younger adults), we repeated the analysis within the overlapping amplitude range of the two age groups (−0.856 to −0.012 µV). Although the amplitude–PLV relationship was reduced in younger adults (r = −0.49), it remained stronger than that observed in older adults (r = −0.06), with a significant age group × CAP amplitude interaction (*p* < 0.001). We also examined the influence of the two younger adults with the largest CAP amplitudes. Excluding these observations reduced the younger-adult correlation from r = −0.85 to r = −0.66, but the relationship remained statistically significant (*p* < 0.001). Together, these analyses indicate that the stronger amplitude–PLV coupling in younger adults is not solely attributable to their broader range of CAP amplitudes or to a small number of extreme observations.

### 2. Residual neural synchrony identifies two physiological AN phenotypes in older adults

To quantify age-related deviations from normative AN physiology, residual neural synchrony was calculated as the difference between the observed PLV and the PLV predicted from the younger-adult CAP amplitude–PLV relationship. Average silhouette width was greatest for a two-cluster solution (0.55 compared with 0.51 and 0.50 for three-and four-cluster solutions, respectively), and gap statistic analysis similarly supported two clusters. Unsupervised k-means clustering was therefore used to define two physiological AN phenotypes characterized by neural synchrony that was either as-expected-or-better or poorer-than-expected relative to CAP amplitude (Fig. 2A).

**Figure 2.**
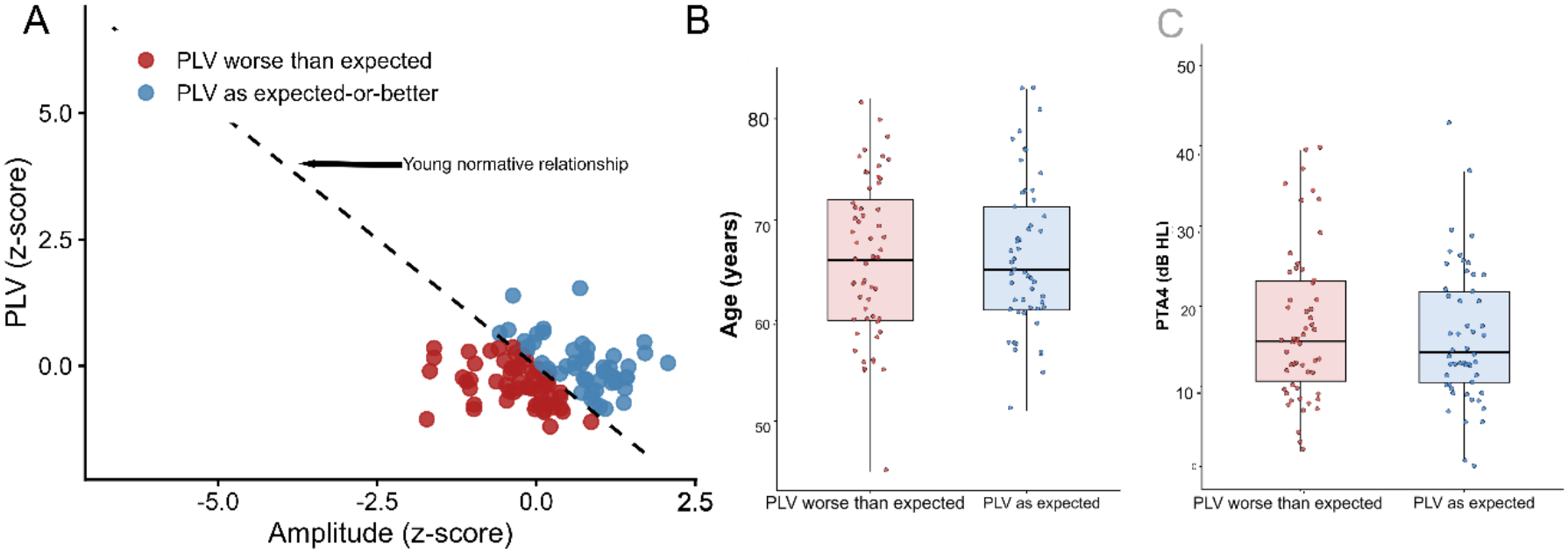
Residual neural synchrony identifies AN phenotypes that are independent of age and hearing thresholds. (A) A normative relationship between CAP amplitude and PLV was established from data from younger adults (dashed line; derived from Figure 1). For each older adult, residual neural synchrony was calculated as the difference between observed PLV and the PLV predicted from the younger-adult normative model. K-means clustering of residual neural synchrony identified two phenotypes: older adults with as-expected-or-better neural synchrony (blue circles) and older adults with poorer-than-expected neural synchrony (red circles). These groups exhibited similar CAP amplitudes but differed in neural synchrony relative to that expected from younger adults. (B) Older adults assigned to the two physiological AN phenotypes did not differ in chronological age (*P* = 0.96). (C) Older adults assigned to the two physiological AN phenotypes did not differ in four-frequency pure-tone average (PTA4; *P* = 0.24).

Older adults in one phenotype exhibited substantially poorer neural synchrony than predicted from the normative model (mean residual = −0.68) and were designated the poorer-than-expected neural synchrony phenotype. The second phenotype was designated the as-expected-or-better neural synchrony phenotype (mean residual = +0.50). Despite marked differences in residual neural synchrony, the two phenotypes did not differ in chronological age (*p* = 0.96), PTA4 (*p* = 0.24), or PTAHF (*p* = 0.22). The phenotypes differed in both CAP amplitude and PLV (both *p* < 0.001), reflecting their different relationships between response magnitude and neural synchrony.

We next evaluated the distribution and stability of the residual-based classification. Gaussian mixture modeling favored a single-component distribution based on the Bayesian information criterion, providing no strong evidence for two discrete underlying populations (Extended data 2-1). Nevertheless, the two-cluster partition showed reasonable stability under bootstrap resampling (median adjusted Rand index = 0.89; 95% bootstrap percentile interval: 0.49–1.00). Classification was largely preserved when residuals were derived using robust regression, with only 2 of 109 older adults changing assignment. Excluding the two younger adults with the largest-magnitude CAP responses from the normative regression resulted in 10 of 109 older adults changing assignment. These findings support the use of the two-group partition as a reasonably stable descriptive classification of continuous physiological variation rather than evidence of discrete biological subtypes.

To assess whether differences in recording quality contributed to the physiological phenotypes, we compared prestimulus measures and accepted trial counts between the two groups. Prestimulus PLV (0.026 vs. 0.028, *p* = 0.23), PPC (0.00001 vs. 0.00003, *p* = 0.74), and RMS voltage (9.05 vs. 8.91 µV, *p* = 0.66) did not differ significantly between phenotypes. Accepted trial counts showed a modest between-phenotype difference (1232 vs. 1158, *p* = 0.050). Among older adults, prestimulus PPC, RMS voltage, and accepted trial count were not significantly associated with evoked PLV (Spearman ρ = 0.152, −0.117, and −0.158, respectively; all *p* > 0.10) or residual neural synchrony (ρ = 0.057, −0.188, and −0.182, respectively; all *p* > 0.05). These findings provide no strong evidence that the physiological phenotype differences are explained by the measured prestimulus recording-quality indices, although modest associations cannot be excluded.

### 3. Residual neural synchrony is associated with auditory nerve diffusion MRI measures and myelin pathology

Compared with the as-expected-or-better phenotype, older adults in the poorer-than-expected phenotype exhibited lower FA (Hedges’ g = −0.54), higher RD (g = 0.62), and higher MD (g = 0.62), corresponding to medium effect sizes. These differences remained significant after controlling for age and audiometric thresholds (PTA4; all *p* < 0.05). Consistent with these group differences, residual neural synchrony was continuously associated with auditory nerve diffusion measures after adjustment for age and PTA4 (Fig. 3A–C). Poorer residual neural synchrony was associated with lower FA (standardized β = 0.235, *p* = 0.047), higher MD (β = −0.244, *p* = 0.040), and higher RD (β = −0.248, *p* = 0.035).

**Figure 3.**
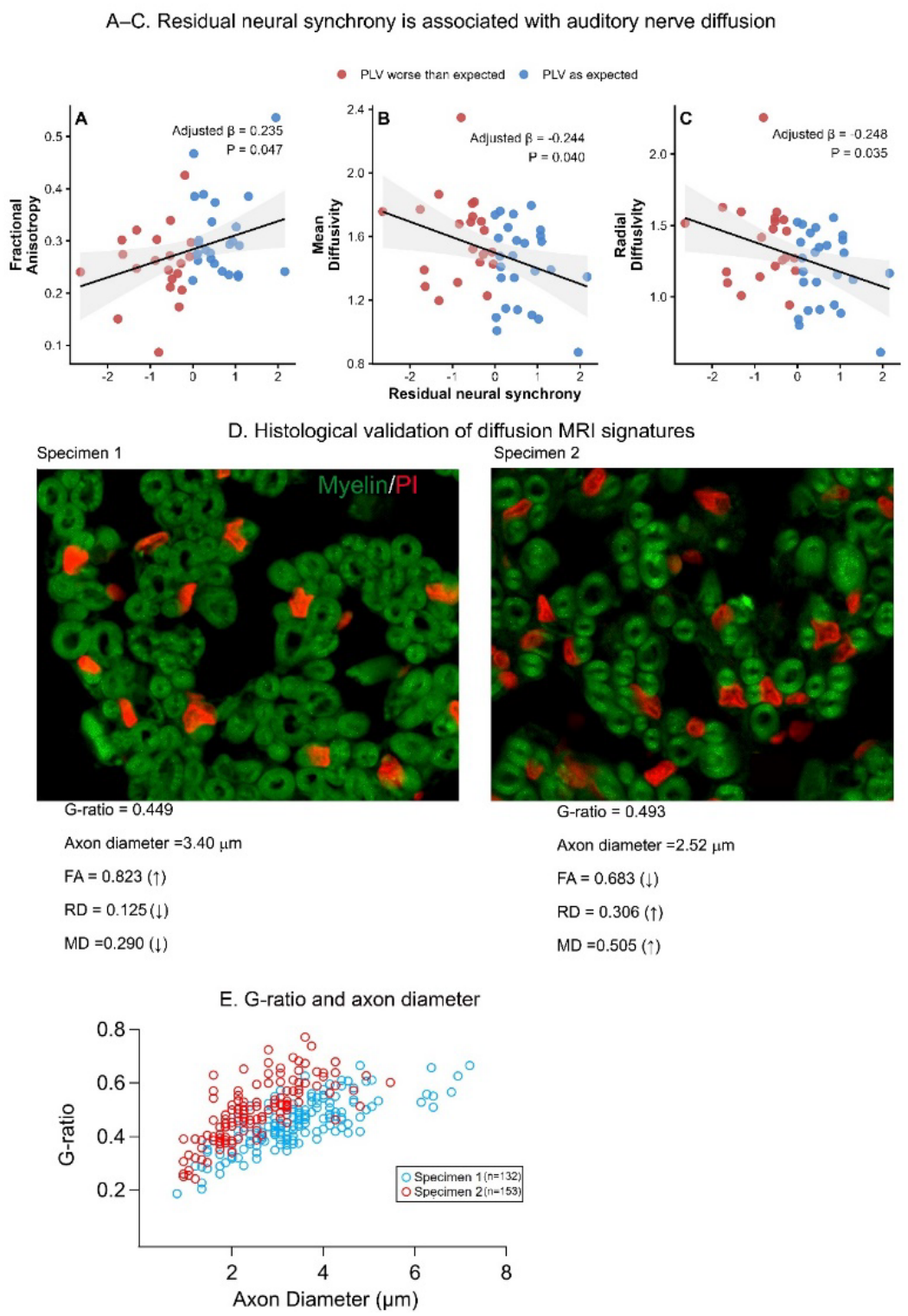
Residual neural synchrony is associated with diffusion MRI measures of AN microstructure and corresponding differences in human AN myelin architecture. (A–C) Residual neural synchrony (observed PLV − PLV predicted from the normative younger-adult CAP amplitude–PLV relationship) was significantly associated with diffusion MRI measures of the AN in older adults after controlling for age and four-frequency pure-tone average (PTA4). Poorer residual neural synchrony was associated with lower fractional anisotropy (FA; A) and higher mean diffusivity (MD; B) and radial diffusivity (RD; C), a pattern of microstructural differences associated with reduced myelin integrity. Points are colored by physiological AN phenotype identified using unsupervised clustering of residual neural synchrony (red, PLV-worse-than-expected; blue, PLV-as-expected-or-better). Regression lines and shaded regions indicate the adjusted linear relationship and 95% confidence interval. (D) Correspondence between diffusion MRI and myelin histology in human temporal bone specimens. FluoroMyelin™ labels the myelin sheath (Myelin, green), and propidium iodide (PI; red) labels cell nuclei. Diffusion MRI measures obtained from the same temporal bone specimens corresponded to differences in histological measures of AN myelin structure, with Specimen 1 exhibiting higher FA and lower MD and RD, together with lower g-ratio and larger myelinated fiber radius, consistent with less myelin degeneration. In contrast, Specimen 2 exhibited lower FA and higher MD and RD, together with higher g-ratio and smaller myelinated fiber radius, consistent with more myelin degeneration. Together, these findings demonstrate an association between residual neural synchrony and AN diffusion MRI measures, with corresponding within-specimen histological evidence linking these diffusion measures to AN myelin integrity. (E) Relationship between g-ratio and axon diameter for myelinated AN fibers from specimen 1 (n = 132) and specimen 2 (n = 153).

Because FA, MD, and RD were strongly correlated, principal component analysis was used to derive a composite measure of auditory nerve microstructure. The first principal component (PC1) explained 92.6% of the variance, with higher scores corresponding to lower FA and higher MD and RD. Among older adults with available electrophysiological and diffusion MRI data (n = 47), residual neural synchrony was associated with PC1 after adjustment for age and PTA4 (standardized β = −0.296, 95% CI [−0.551, −0.040], *p* = 0.034). In contrast, the adjusted difference in PC1 between physiological phenotypes was not statistically significant (*p* = 0.141).

To determine whether residual neural synchrony provided information beyond the individual electrophysiological measures, we compared associations with auditory nerve diffusion measures. CAP amplitude was not significantly correlated with any diffusion measure (all *p* > 0.19), whereas raw PLV and residual neural synchrony were both associated with FA, MD, RD, and AD (all unadjusted *p* < 0.05), with generally comparable association strengths. In complete-case models of PC1 (n = 47), age and PTA4 explained 25.7% of the variance. Adding CAP amplitude increased R² to 0.30 (*p* = 0.141), raw PLV to 0.32 (*p* = 0.049), and residual neural synchrony to 0.34 (*p* = 0.024). Although residual synchrony showed the largest incremental association with PC1, adding CAP amplitude to a model already containing raw PLV did not significantly improve fit (*p* = 0.160), indicating that residual PLV was not demonstrably superior to raw PLV.

The association between residual neural synchrony and PC1 remained significant when residuals were derived using robust regression (β = −0.286, *p* = 0.028) or after excluding the two younger adults with the largest-magnitude CAP responses from the normative model (β = −0.304, *p* = 0.020). This association was also robust to additional adjustment for ROI size (*p* = 0.039; ROI size: *p* = 0.783) and recording-quality measures, including prestimulus PPC, baseline RMS amplitude, and accepted trial count (β = −0.287, *p* = 0.045).

To examine whether the diffusion measures observed in living participants corresponded with auditory nerve myelin pathology, we compared diffusion MRI and histology from two postmortem human temporal bone specimens. Specimen 1 (50s years), characterized by higher FA (0.823) and lower RD (0.125) and MD (0.290), exhibited a lower mean g-ratio (0.449) and larger mean axon diameter (3.40 µm) than specimen 2 (80s years), which exhibited lower FA (0.683), higher RD (0.306) and MD (0.505), a higher mean g-ratio (0.493), and smaller mean axon diameter (2.52 µm; Fig. 3D). Quantitative histological analysis included 132 myelinated fibers from specimen 1 and 153 from specimen 2. Examination of g-ratio as a function of axon diameter (Fig. 3E; Extended Data Figure 3-1) further demonstrated differences in myelin structure and axon caliber between specimens. These observations provide anatomical context for the corresponding diffusion MRI differences, while recognizing that myelin and axonal alterations may coexist. Although limited to two specimens, these findings provide preliminary histological support for the interpretation that variation in auditory nerve diffusion measures may reflect differences in myelin integrity. Together with the physiological–diffusion associations observed in living participants, these findings support a potential contribution of myelin degeneration to altered auditory nerve synchrony.

### 4. Physiological AN phenotype is associated with the agreement between audiometric thresholds and self-reported hearing difficulty

To determine whether physiological AN phenotype was associated with self-reported hearing difficulty beyond that explained by audiometric thresholds, we used hierarchical linear regression to predict Revised Hearing Handicap Inventory (RHHI) scores. Although RHHI data were available for 74 older adults, complete data for the physiological and audiometric predictors were available for 70 older adults included in these analyses. Audiometric thresholds (PTA4) accounted for 21.5% of the variance in RHHI scores (R² = 0.215, *p* < 0.001). Adding physiological AN phenotype explained an additional 2.8% of the variance (ΔR² = 0.028), although this improvement was not statistically significant (*p* = 0.157). However, adding the PTA4 × phenotype interaction significantly improved model fit, explaining an additional 6.0% of the variance (ΔR² = 0.060, *p* = 0.019). The final model explained 30.2% of the variance in self-reported hearing difficulty (R² = 0.302). As shown in Fig. 4 and Table 1, the relationship between audiometric thresholds and RHHI scores differed according to physiological AN phenotype. Among older adults with poorer-than-expected neural synchrony, RHHI scores increased more steeply with increasing audiometric thresholds than among those with as-expected-or-better neural synchrony. Thus, physiological phenotype was associated with differences in how audiometric hearing loss related to self-reported hearing difficulty.

**Figure 4.**
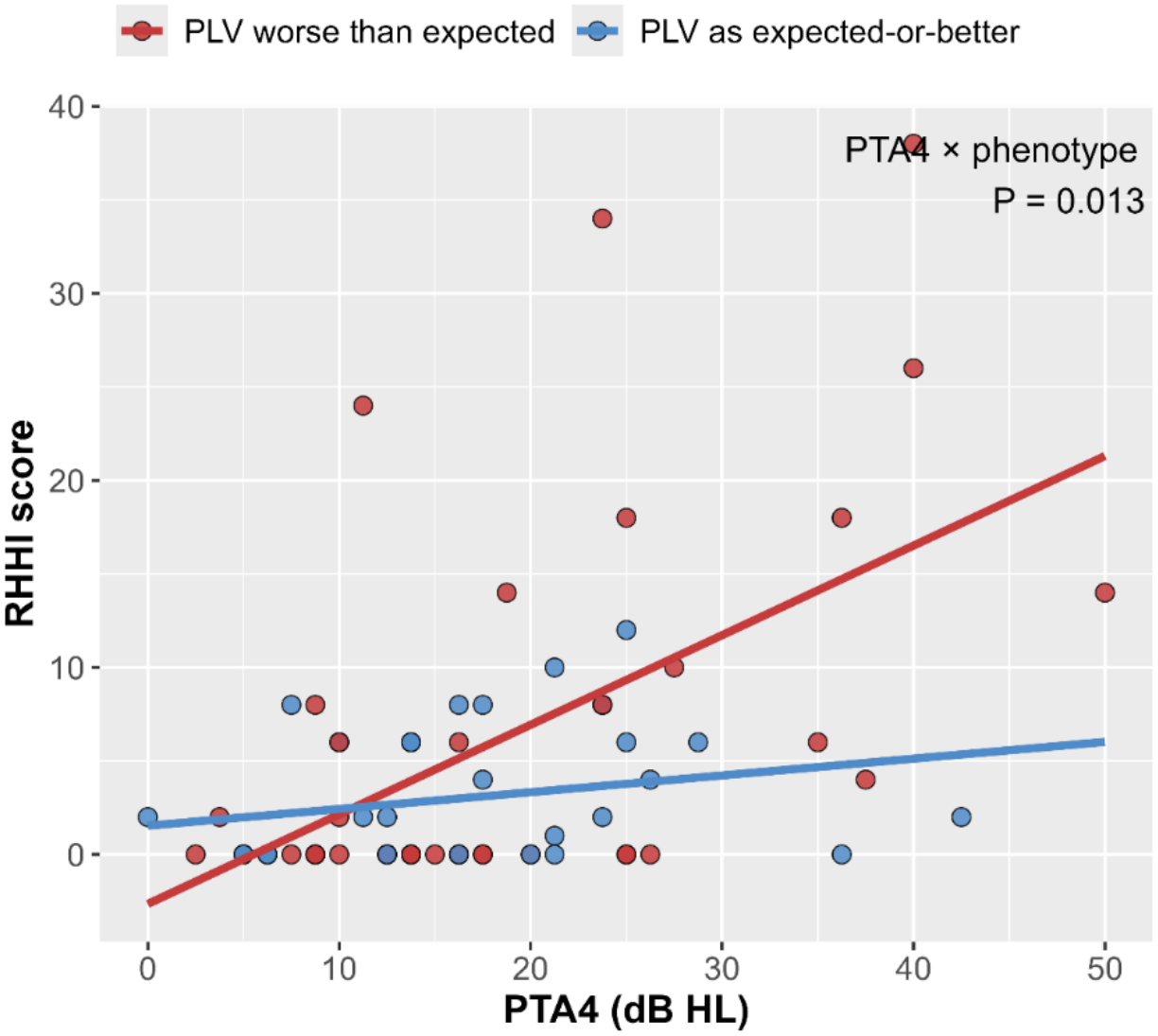
Physiological AN phenotype is associated with the agreement between audiometric thresholds and self-reported hearing difficulty. RHHI scores are plotted as a function of PTA4 for older adults with PLV-worse-than-expected neural synchrony (red circles) and PLV-as-expected neural synchrony (blue circles). The significant PTA4 × phenotype interaction (*p* = 0.019) indicates that the agreement between audiometric thresholds and self-reported hearing difficulty differed according to AN phenotype.

**Table 1:** RHHI, PTA4, and Physiologic auditory nerve phenotype.

| **Model** | **Predictors** | **R<sup>2</sup>** | **ΔR<sup>2</sup>** | **F change** | **P** |
|---|---|---|---|---|---|
| 1 | PTA4 | 0.215 | — | — | <0.001 |
| 2 | + Physiological phenotype | 0.243 | 0.028 | 2.05 | 0.157 |
| 3 | + PTA4 ×<br>Physiological phenotype | **0.302** | **0.06** | **4.42** | **0.019** |

**Table 2.** Participant characteristics and availability of study measures.

| **Characteristic** | **Younger adults** | **Older adults** |
|---|---|---|
| Participants | 69 | 113 |
| Age, years (range) | 18–30 | 54–83 |
| Age, years (mean ± SD) | 24.1 ± 3.1 | 66.3 ± 7.4 |
| Sex (CAP-complete; F / M) | 48 / 19 | 74 / 35 |
| PTA4, dB HL (mean ± SD) | 2.3 ± 4.0 | 17.2 ± 9.5 |
| PTAHF, dB HL (mean ± SD) | 3.1 ± 4.2 | 29.0 ± 15.6 |
| CAP amplitude and PLV available | 67 | 109 |
| Diffusion MRI available | 18 | 47 |
| RHHI available | 31 | 74 |
| Both diffusion MRI and RHHI | 11 | 34 |

To examine whether residual neural synchrony provided information beyond the individual physiological measures, we compared models incorporating interactions between PTA4 and CAP amplitude, raw PLV, or residual neural synchrony in the same 70 older adults. The model incorporating the PTA4 × residual neural synchrony interaction explained 34.0% of the variance in RHHI scores, compared with 28.6% for raw PLV and 28.0% for CAP amplitude. The interaction between PTA4 and residual neural synchrony was statistically significant (*p* = 0.002). Although the residual-based model explained numerically more variance than models incorporating either physiological measure independently, these comparisons do not establish statistically superior performance. Nevertheless, the findings support the potential behavioral relevance of evaluating neural synchrony relative to AN response amplitude.

Sex-related analyses. Sex distribution did not differ significantly between the poorer-than-expected and as-expected-or-better PLV phenotypes (35 female/22 male and 39 female/13 male, respectively; Fisher’s exact test, *p* = 0.153). Importantly, the association between residual PLV and auditory nerve diffusion PC1 remained significant after adjusting for age, hearing thresholds (PTA4), and sex (HC3 robust *p* = 0.029). Similarly, the relationship between CAP amplitude and PLV in younger adults remained significant after accounting for sex (*p* = 0.003).

## Discussion

Deviations from the normative relationship between AN response amplitude and neural synchrony (PLV) reveal physiologically meaningful variation in AN aging. An individual with small CAP amplitude and low PLV may exhibit substantial neural dysfunction, but these measures provide limited mechanistic insight. In contrast, PLV that is disproportionately low relative to CAP amplitude suggests that temporal synchrony is more impaired than expected for the magnitude of the neural response, potentially implicating processes such as myelin degeneration. Poorer residual synchrony was associated with diffusion MRI measures of AN microstructure, with illustrative histological support from human temporal bones. The relationship between audiometric thresholds and self-reported hearing difficulty varied by physiological phenotype, with a similar association for continuous residual synchrony.

Previous studies of AN aging have focused on reduced response amplitude and, to a lesser extent, prolonged latencies (Burkard and Sims, 2001; Konrad-Martin et al., 2012; Mehraei et al., 2016; Bramhall et al., 2017). Fewer studies have examined AN synchrony, and amplitude and PLV have generally been interpreted independently. In younger adults with normal hearing, response amplitude and neural synchrony were tightly coupled, whereas this association was markedly weakened in aging. The proportion of older adults exhibiting lower-than-expected neural synchrony suggests that deviations from the normative relationship between response amplitude and synchrony may be relatively common in aging. Although negative residuals do not necessarily indicate pathology, this approach may help estimate the prevalence of meaningful AN dysfunction once normative limits and pathological validation are established. This parallels normative modeling elsewhere in neuroscience, where deviations from expected biological relationships reveal heterogeneity obscured by group-based analyses (Wolfers et al., 2018; Marquand et al., 2019). Although the Gaussian mixture analysis favored a single residual distribution, indicating no strong evidence for two discrete underlying populations, variation along this continuous physiological dimension was associated with AN microstructure and self-reported hearing difficulty. The two-group partition summarizes this variation without establishing separate disease entities.

The normative approach may be valuable across stages of neural dysfunction. For example, an individual with relatively large CAP amplitudes could retain higher absolute PLV than an older adult with reduced AN input yet exhibit poorer synchrony than expected for their response amplitude. Such deviations could reveal emerging dysfunction before absolute PLV becomes abnormal, although their relationship to early myelin pathology requires longitudinal validation. Residual PLV showed a numerically stronger association with AN microstructure than raw PLV, without established predictive superiority. Importantly, residual PLV explained more variance in self-reported hearing difficulty than either raw PLV or CAP amplitude alone, supporting the behavioral relevance of evaluating synchrony relative to response magnitude.

Three complementary lines of evidence support an association between residual neural synchrony and AN myelin integrity. First, poorer residual neural synchrony was associated with lower FA and higher RD and MD, a pattern of diffusion MRI changes that has been associated with reduced myelin integrity in previous histological validation studies (Song et al., 2002; Jelescu and Budde, 2017; Lazari and Lipp, 2021). The strong loading of FA, RD, and MD onto one principal component suggests a common dimension of AN microstructure. This convergent pattern is particularly relevant in the relatively coherent fiber architecture of peripheral nerves, where FA and RD have been associated with electrophysiological measures sensitive to myelin integrity. Although not myelin-specific, these convergent measures support a myelin-associated interpretation of AN microstructural differences (Heckel et al., 2015). Second, these diffusion MRI differences were consistent with myelin structural differences in two human temporal bone specimens. The specimen with higher FA and lower RD and MD exhibited a lower g-ratio and larger myelinated axon diameter, whereas the specimen with lower FA and higher RD and MD exhibited a higher g-ratio and smaller myelinated axon diameter. Third, these findings are consistent with our previous studies in mouse models demonstrating that AN myelin and nodal organization are closely linked to neural synchrony (Panganiban et al., 2022). Although no single approach provides definitive evidence that AN myelin degeneration is the sole mechanism underlying reduced neural synchrony, these complementary findings implicate AN myelin as a potential contributor to the physiological variation observed in older adults.

Importantly, physiological and structural measures do not necessarily sample identical portions of the AN. The peripheral AN is myelinated by Schwann cells, whereas the central portion is myelinated by oligodendrocytes, with the glial transition zone (GTZ) forming a specialized interface between these domains (Payne et al., 2026). The TM-recorded CAP reflects the synchronized population response of the AN. Although action potentials are thought to initiate near the first heminode, the compound field potential recorded at the TM reflects summed neural activity and cannot be localized to a discrete segment of the AN. Similarly, the spatial resolution of the present diffusion MRI does not permit reliable separation of peripheral, GTZ, and central AN compartments. Our MRI regions were defined conservatively to include only portions of the AN identified with confidence rather than imposing fixed anatomical boundaries corresponding to specific myelin compartments. These measures may reflect related aspects of AN structure and function, although their anatomical overlap remains uncertain. Importantly, available anatomical evidence indicates that age-related myelin pathology is not restricted to a single AN compartment, with abnormalities reported in peripheral and central regions and emerging evidence that the GTZ may be particularly vulnerable to age-related glial cell dysfunction and altered immune activity (Payne et al., 2026). However, the present methods cannot determine whether myelin abnormalities are quantitatively coupled across these compartments within individuals or localize the structural substrate underlying reduced neural synchrony to a specific portion of the AN. Despite these limitations, myelin remains a compelling therapeutic target because, unlike permanent neuronal loss, myelin retains the capacity for self-repair and remodeling (Ettle et al., 2016; Chapman and Hill, 2020). Pharmacological strategies promoting remyelination are advancing across neurology, underscoring the need for biomarkers capable of identifying myelin degeneration and monitoring therapeutic response. These findings provide a framework for developing *in vivo* biomarkers linking neural synchrony to diffusion MRI measures of auditory nerve microstructure associated with myelin integrity. Perhaps most importantly, the physiological AN phenotypes identified here linked AN physiology to patient experience. The relationship between audiometric thresholds and self-reported hearing difficulty differed according to physiological phenotype, with hearing difficulty increasing more steeply as thresholds worsened among older adults with poorer-than-expected neural synchrony. Continuous residual neural synchrony showed a similar association, suggesting that individual deviations from normative AN physiology may help explain variability in communication difficulties not captured by audiometric thresholds alone. These findings support the potential clinical relevance of characterizing neural synchrony relative to AN response amplitude, although further studies are needed to establish its incremental predictive value and clinical utility.

Several limitations should be considered. Although the diffusion MRI findings were supported by corresponding histological differences in human temporal bone specimens, only two specimens from two donors were examined. Detailed premortem hearing histories were unavailable; however, the purpose of the MRI–histology comparison was not to establish the etiology or functional consequences of AN pathology, but to determine whether differences in diffusion MRI corresponded to differences in underlying nerve microstructure. Histological measures of myelin and axonal integrity can be characterized directly, regardless of whether the observed pathology arose from aging, hearing loss, noise exposure, or other causes. Thus, the absence of premortem auditory information does not preclude evaluation of MRI–histology correspondence. Larger studies incorporating quantitative measures of both myelin and axonal degeneration will be necessary to establish the specificity and reproducibility of these MRI–histology relationships. Approaches such as whole-nerve evaluation using a tissue clearing approach (Payne et al., 2026) may further strengthen this validation by providing more comprehensive three-dimensional characterization of AN architecture and improving anatomical correspondence between histological and MRI measures. The diffusion MRI analyses included a relatively small subset of older adults, and the observed associations require replication in larger, independent samples. Although the converging diffusion MRI, temporal bone, and prior findings in mouse models implicate myelin degeneration as an important contributor to poorer-than-expected neural synchrony, reduced synchrony is unlikely to be specific to myelin pathology. Other forms of AN pathology, including synaptic loss and disruption of nodal or heminodal organization, may also alter the relationship between response amplitude and neural synchrony. The present study used an unsupervised physiological classification designed to determine whether biologically meaningful phenotypes emerge directly from human data. However, the residual distribution did not provide strong evidence for discrete underlying populations, and the two-group classification should be considered descriptive rather than diagnostic. An important next step will be to extend this framework using supervised approaches trained on animal models with known underlying AN pathology. Such models could determine the extent to which physiological profiles distinguish specific mechanisms, including myelin degeneration and synaptic loss, while retaining the physiological interpretability established here.

More broadly, these findings demonstrate that normative physiological relationships can characterize biologically relevant physiological heterogeneity not captured by current clinical tests. As therapies targeting myelin repair continue to advance, this framework provides a foundation for developing physiological biomarkers of age-related AN dysfunction, with the longer-term goal of identifying underlying mechanisms, informing patient stratification, and monitoring responses to targeted interventions.

## Data and Code Availability

De-identified participant-level data underlying the findings of this study, including demographic, audiometric, electrophysiological, diffusion MRI, and self-reported hearing measures, will be deposited in a research data repository upon publication, consistent with NIH Data Management and Sharing requirements. Because data were collected across multiple protocols over approximately 15 years, the deposited dataset will include participants for whom data sharing is permitted under the applicable informed consent and institutional approvals. The AN CAP processing pipeline is publicly available through co-author McClaskey’s GitHub repository. Diffusion MRI preprocessing and tensor estimation were performed using the publicly available PyDesigner software (Dhiman et al., 2024). Custom code implementing the residual-based approach to identifying AN myelin degeneration is currently subject to intellectual property development and is not publicly available.

## Data Availability

All data produced in the present study are available upon reasonable request to the authors

## Acknowledgments

This work was supported by the National Institutes of Health/National Institute on Deafness and Other Communication Disorders under Grant Award Numbers R01 DC022765, R01 DC017619, and P50 DC000422

## Author Contributions

K.C.H. conceived and designed the study, processed and analyzed the data, and wrote the manuscript. C.M.M., J.W.D., S.A.P., A.L., C.H., J.R.D, and H.L. contributed to data collection and/or data analysis, and/or critically revised and edited the manuscript. All authors reviewed and approved the final manuscript.

---

**Extended Data Figure 2-1.**
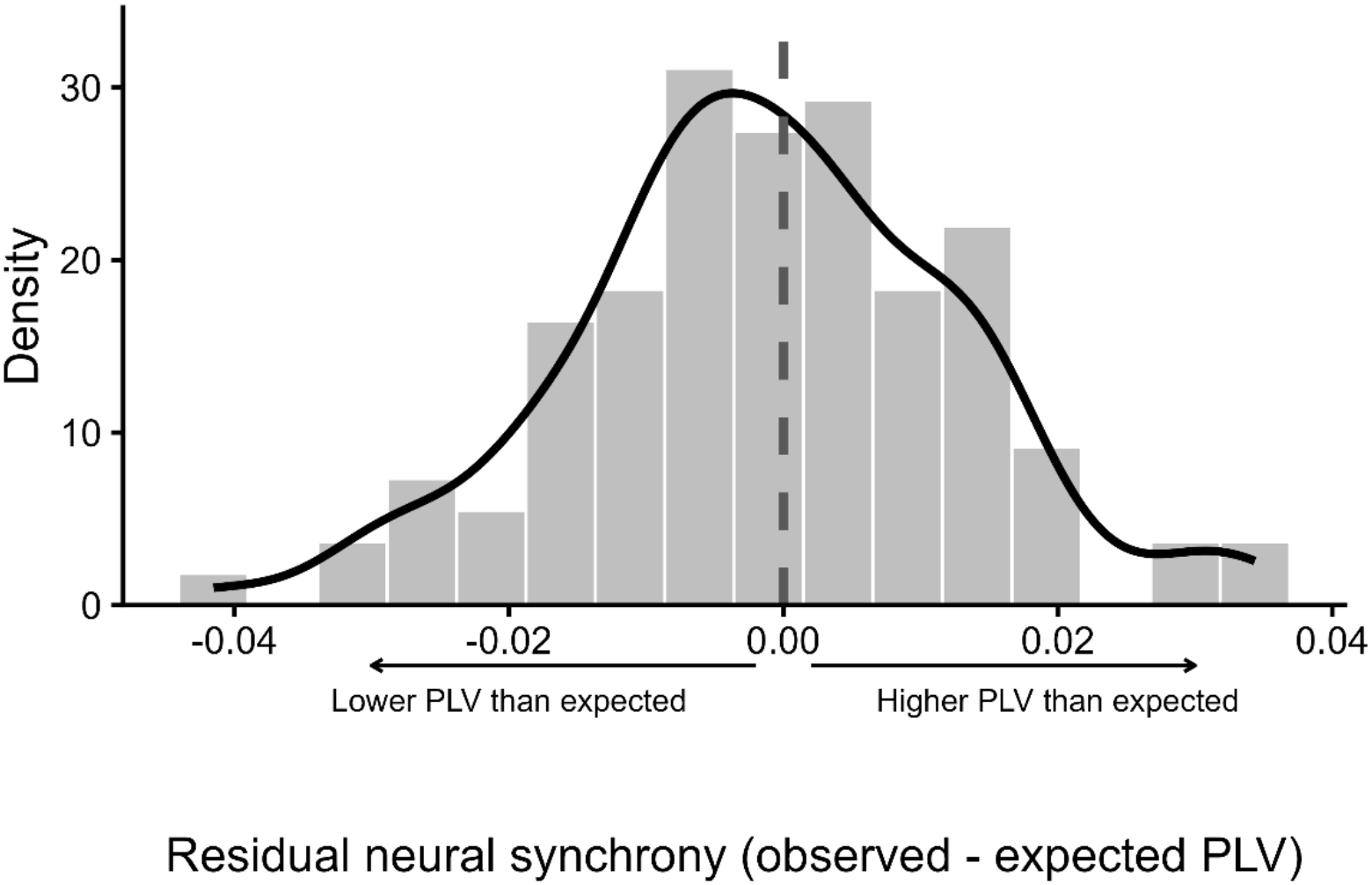
Distribution of residual neural synchrony in older adults. Histogram and kernel density estimate of residual neural synchrony, calculated as observed minus expected PLV based on the normative relationship between CAP amplitude and PLV in younger adults. The dashed vertical line indicates zero residual synchrony, corresponding to the expected PLV for a given CAP amplitude. Negative residuals (left of zero) indicate poorer-than-expected PLV, whereas positive residuals (right of zero) indicate as-expected-or-better neural synchrony phenotypes, as illustrated by the directional arrows. The distribution is continuous, without an obvious separation into discrete subgroups, supporting the interpretation of physiological phenotypes as descriptive divisions of continuous variation rather than distinct biological subtypes.

**Extended Data Figure 3-1.**
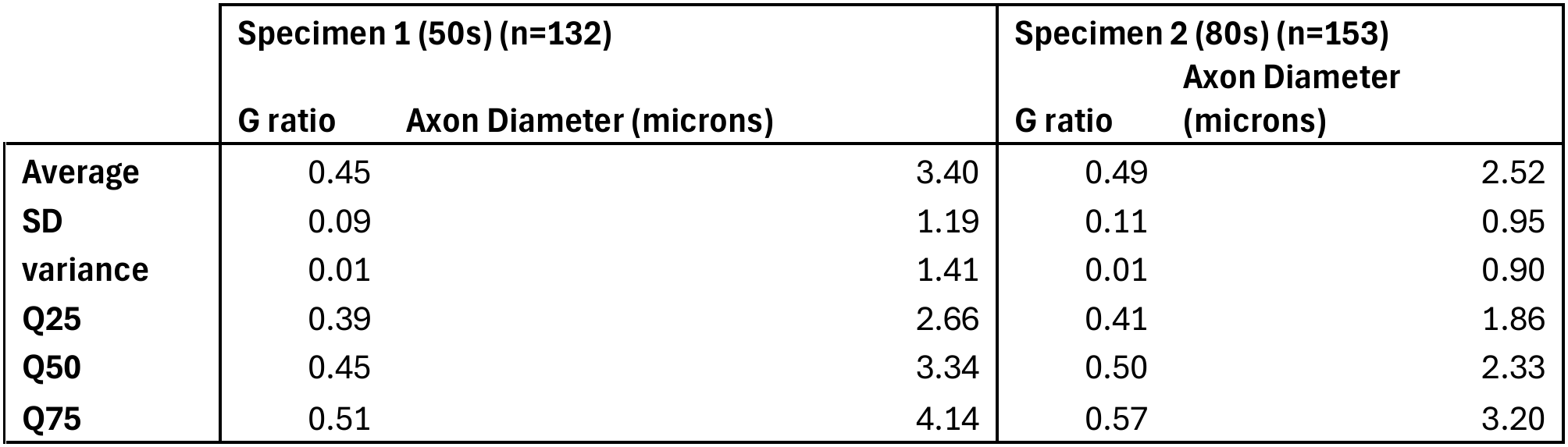
Human temporal bone_G-ratio and axon diameter

## References

Bonetto G, Belin D, Káradóttir RT (2021) Myelin: a gatekeeper of activity-dependent circuit plasticity? Science 374:eaba6905.

Bonetto G, Kamen Y, Evans KA, Káradóttir RT (2020) Unraveling Myelin Plasticity. Frontiers in cellular neuroscience 14:156.

Bramhall NF, Konrad-Martin D, McMillan GP, Griest SE (2017) Auditory Brainstem Response Altered in Humans With Noise Exposure Despite Normal Outer Hair Cell Function. Ear and hearing 38:e1–e12.

Burkard RF, Sims D (2001) The human auditory brainstem response to high click rates: aging effects. American journal of audiology 10:53–61.

Cassarly C, Matthews LJ, Simpson AN, Dubno JR (2020) The Revised Hearing Handicap Inventory and Screening Tool Based on Psychometric Reevaluation of the Hearing Handicap Inventories for the Elderly and Adults. Ear Hear 41:95–105.

Chapman TW, Hill RA (2020) Myelin plasticity in adulthood and aging. Neurosci Lett 715:134645.

Dhiman S, Hickey R, Thorn K, Moss H, McKinnon E, Adisetiyo V, Ades-Aron B, Jensen J, Benitez A (2024) PyDesigner v1.0: A Pythonic Implementation of the DESIGNER Pipeline for Diffusion Magnetic Resonance Imaging. Journal of Visualized Experiments.

Dias JW, McClaskey CM, Alvey AP, Lawson A, Matthews LJ, Dubno JR, Harris KC (2024) Effects of age and noise exposure history on auditory nerve response amplitudes: A systematic review, study, and meta-analysis. Hear Res 447:109010.

Dong S, Mulders WH, Rodger J, Woo S, Robertson D (2010) Acoustic trauma evokes hyperactivity and changes in gene expression in guinea-pig auditory brainstem. Eur J Neurosci 31:1616–1628.

Engineer ND, Riley JR, Seale JD, Vrana WA, Shetake JA, Sudanagunta SP, Borland MS, Kilgard MP (2011) Reversing pathological neural activity using targeted plasticity. Nature 470:101–104.

Ettle B, Schlachetzki JCM, Winkler J (2016) Oligodendroglia and Myelin in Neurodegenerative Diseases: More Than Just Bystanders? Mol Neurobiol 53:3046–3062.

Franklin RJM, Ffrench-Constant C (2017) Regenerating CNS myelin – from mechanisms to experimental medicines. Nat Rev Neurosci 18:753–769.

Gates GA, Mills JH (2005) Presbycusis. Lancet 366:1111–1120.

Harris KC, Vaden KI, Jr., McClaskey CM, Dias JW, Dubno JR (2018) Complementary metrics of human auditory nerve function derived from compound action potentials. Journal of neurophysiology 119:1019–1028.

Harris KC, Dias JW, McClaskey CM, Rumschlag J, Prisciandaro J, Dubno JR (2022) Afferent loss, GABA, and Central Gain in older adults: Associations with speech recognition in noise. J Neurosci.

Harris KC, Ahlstrom JB, Dias JW, Kerouac LB, McClaskey CM, Dubno JR, Eckert MA (2021) Neural Presbyacusis in Humans Inferred from Age-Related Differences in Auditory Nerve Function and Structure. J Neurosci 41:10293–10304.

Hartigan JA, Wong MA (1979) A K-Means Clustering Algorithm. Journal of the Royal Statistical Society: Series C (Applied Statistics) 28:100–108.

Heckel A, Weiler M, Xia A, Ruetters M, Pham M, Bendszus M, Heiland S, Baeumer P (2015) Peripheral Nerve Diffusion Tensor Imaging: Assessment of Axon and Myelin Sheath Integrity. PLoS One 10:e0130833.

Hickman TT, Hashimoto K, Liberman LD, Liberman MC (2021) Cochlear Synaptic Degeneration and Regeneration After Noise: Effects of Age and Neuronal Subgroup. Frontiers in cellular neuroscience 15:684706.

Jelescu IO, Budde MD (2017) Design and Validation of Diffusion MRI Models of White Matter. Frontiers in Physics Volume 5–2017.

Konrad-Martin D, Dille MF, McMillan G, Griest S, McDermott D, Fausti SA, Austin DF (2012) Age-related changes in the auditory brainstem response. J Am Acad Audiol 23:18–75.

Kotak VC, Fujisawa S, Lee FA, Karthikeyan O, Aoki C, Sanes DH (2005) Hearing loss raises excitability in the auditory cortex. J Neurosci 25:3908–3918.

Kraus KS, Ding D, Jiang H, Lobarinas E, Sun W, Salvi RJ (2011) Relationship between noise-induced hearing-loss, persistent tinnitus and growth-associated protein-43 expression in the rat cochlear nucleus: does synaptic plasticity in ventral cochlear nucleus suppress tinnitus? Neuroscience 194:309–325.

Kujawa SG, Liberman MC (2015) Synaptopathy in the noise-exposed and aging cochlea: Primary neural degeneration in acquired sensorineural hearing loss. Hearing research 330:191–199.

Lazari A, Lipp I (2021) Can MRI measure myelin? Systematic review, qualitative assessment, and meta-analysis of studies validating microstructural imaging with myelin histology. Neuroimage 230:117744.

Makary CA, Shin J, Kujawa SG, Liberman MC, Merchant SN (2011) Age-related primary cochlear neuronal degeneration in human temporal bones. J Assoc Res Otolaryngol 12:711–717.

Marquand AF, Kia SM, Zabihi M, Wolfers T, Buitelaar JK, Beckmann CF (2019) Conceptualizing mental disorders as deviations from normative functioning. Molecular Psychiatry 24:1415–1424.

Mehraei G, Hickox AE, Bharadwaj HM, Goldberg H, Verhulst S, Liberman MC, Shinn-Cunningham BG (2016) Auditory Brainstem Response Latency in Noise as a Marker of Cochlear Synaptopathy. J Neurosci 36:3755–3764.

Panganiban CH, Barth JL, Tan J, Noble KV, McClaskey CM, Howard BA, Jafri SH, Dias JW, Harris KC, Lang H (2022) Two distinct types of nodes of Ranvier support auditory nerve function in the mouse cochlea. Glia 70:768–791.

Panganiban CH, Barth JL, Darbelli L, Xing Y, Zhang J, Li H, Noble KV, Liu T, Brown LN, Schulte BA, Richard S, Lang H (2018) Noise-Induced Dysregulation of Ǫuaking RNA Binding Proteins Contributes to Auditory Nerve Demyelination and Hearing Loss. J Neurosci 38:2551–2568.

Parthasarathy A, Bartlett EL, Kujawa SG (2019) Age-related Changes in Neural Coding of Envelope Cues: Peripheral Declines and Central Compensation. Neuroscience 407:21–31.

Payne SA, Anderson HR, Brown IM, Chai J, Chen P, Yao H, Barth JL, Lang H (2026) The auditory nerve glial transition zone is a focal site of age-related immune-myelin interactions. Front Immunol 17:1757234.

Popescu MV, Polley DB (2010) Monaural deprivation disrupts development of binaural selectivity in auditory midbrain and cortex. Neuron 65:718–731.

Rumschlag JA, McClaskey CM, Dias JW, Kerouac LB, Noble KV, Panganiban C, Lang H, Harris KC (2022) Age-related central gain with degraded neural synchrony in the auditory brainstem of mice and humans. Neurobiol Aging 115:50–59.

Salvi R, Sun W, Ding D, Chen GD, Lobarinas E, Wang J, Radziwon K, Auerbach BD (2016) Inner Hair Cell Loss Disrupts Hearing and Cochlear Function Leading to Sensory Deprivation and Enhanced Central Auditory Gain. Front Neurosci 10:621.

Sanes DH, Bao S (2009) Tuning up the developing auditory CNS. Curr Opin Neurobiol 19:188–199.

Schmiedt RA, Okamura HO, Lang H, Schulte BA (2002) Ouabain application to the round window of the gerbil cochlea: a model of auditory neuropathy and apoptosis. J Assoc Res Otolaryngol 3:223–233.

Seki S, Eggermont JJ (2003) Changes in spontaneous firing rate and neural synchrony in cat primary auditory cortex after localized tone-induced hearing loss. Hear Res 180:28–38.

Song SK, Sun SW, Ramsbottom MJ, Chang C, Russell J, Cross AH (2002) Dysmyelination revealed through MRI as increased radial (but unchanged axial) diffusion of water. Neuroimage 17:1429–1436.

Suthakar K, Liberman MC (2021) Auditory-nerve responses in mice with noise-induced cochlear synaptopathy. J Neurophysiol 126:2027–2038.

Verdi S, Kia SM, Yong KXX, Tosun D, Schott JM, Marquand AF, Cole JH (2023) Revealing Individual Neuroanatomical Heterogeneity in Alzheimer Disease Using Neuroanatomical Normative Modeling. Neurology 100:e2442–e2453.

Verdi S, Rutherford S, Fraza C, Tosun D, Altmann A, Raket LL, Schott JM, Marquand AF, Cole JH (2024) Personalizing progressive changes to brain structure in Alzheimer’s disease using normative modeling. Alzheimers Dement 20:6998–7012.

Wan G, Corfas G (2017) Transient auditory nerve demyelination as a new mechanism for hidden hearing loss. Nat Commun 8:14487.

Wolfers T, Doan NT, Kaufmann T, Alnæs D, Moberget T, Agartz I, Buitelaar JK, Ueland T, Melle I, Franke B, Andreassen OA, Beckmann CF, Westlye LT, Marquand AF (2018) Mapping the Heterogeneous Phenotype of Schizophrenia and Bipolar Disorder Using Normative Models. JAMA Psychiatry 75:1146–1155.

Wu PZ, Liberman LD, Bennett K, de Gruttola V, O’Malley JT, Liberman MC (2019) Primary Neural Degeneration in the Human Cochlea: Evidence for Hidden Hearing Loss in the Aging Ear. Neuroscience 407:8–20.

Xing Y, Samuvel DJ, Stevens SM, Dubno JR, Schulte BA, Lang H (2012) Age-related changes of myelin basic protein in mouse and human auditory nerve. PLoS One 7:e34500.

